## Supplementary figures and images for "SplitStrains, a tool to identify and separate mixed *Mycobacterium tuberculosis* infections from WGS data"

### Supplementary Figure 1

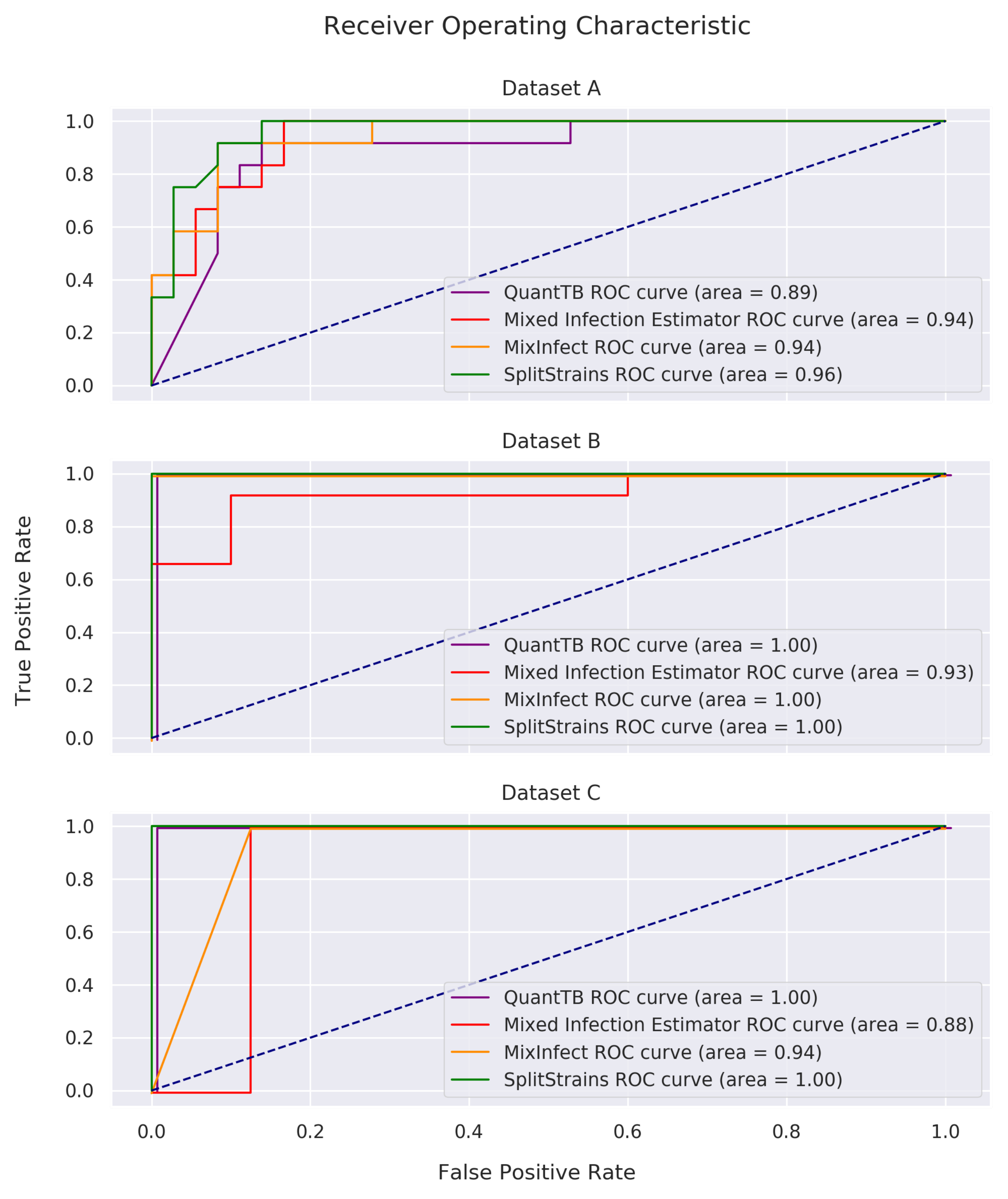
